# A rare congenital anemia is caused by mutations in the centralspindlin complex

**DOI:** 10.1101/2021.04.07.21254990

**Authors:** Sandeep N. Wontakal, Mishan Britto, Sarah Tannenbaum, Benjamin H. Durham, Margaret T. Lee, Masanori Mishima

## Abstract

Congenital dyserythropoietic anemias (CDAs) are rare disorders characterized by morphologic abnormalities of erythroid precursors leading to ineffective erythropoiesis. CDA type III (CDAIII), characterized by erythroblast multinucleation, represents the rarest form with only ∼60 patients described in the literature. Previous work, studying two independent families, identified a causative dominant missense mutation in *KIF23*, which encodes for the kinesin MKLP1. Here, we describe a sporadic CDAIII case associated with compound heterozygous variants in *RACGAP1*, a gene not previously associated with any disease. *RACGAP1* encodes CYK4, a GTPase activating protein (GAP) for Rho-family GTPases, which interacts with MKLP1 to form the centralspindlin complex. Functional assays show these *RACGAP1* variants cause cytokinesis defects due, at least in part, to altering the substrate specificities of the GAP activity of CYK4. These findings provide novel insights into the structural determinants of the GAP activity and demonstrates that cytokinesis failure due to centralspindlin defects leads to CDAIII. Our findings highlight the importance of viewing diseases as malfunctions of common biological pathways/complexes and suggests that next-generation sequencing analysis pipelines should integrate a systems approach in order to identify such functionally related variants.

## Introduction

Congenital dyserythropoietic anemias (CDAs) are a heterogenous group of rare inherited disorders characterized by ineffective erythropoiesis leading to anemia. CDAs were classically distinguished into three major types based on erythroblast morphology in the bone marrow; a fourth type emerged that encompassing atypical morphologies (1, 2). The genetic etiology of these disorders include mutations in *CDAN1* or *C15ORF41* for CDAI (3, 4), *SEC23B* for CDAII (5, 6), *KIF23* for CDAIII (7), and *KLF1, GATA1, ALAS2*, or *VPS4A* for some of the CDA variants (8–11). CDAIII is the rarest form with ∼60 patients described.

The characteristic abnormality seen in CDAIII is multinucleated erythroblasts that are reminiscent of cells undergoing endoreplication (i.e., DNA replication without cell division). Studies of two independent CDAIII families identified a dominant missense mutation in *KIF23* (c.2747C>G; p.P916R) that segregated with disease (7). *KIF23* encodes MKLP1, the kinesin subunit of centralspindlin, which is essential for proper cytokinesis (12). Functional studies of the p.P916R mutation demonstrated failure of proper cytokinesis resulting in multinucleated cells, the hallmark of CDAIII (7).

The original CDAIII families showed an autosomal dominant inheritance pattern (13–15). However, a few sporadic cases were reported with variability in clinical presentation that seem to have an autosomal recessive pattern of inheritance, suggesting genetic heterogeneity (1, 2). To date, no genetic cause has been identified in such autosomal recessive cases. Here, we report a sporadic case of CDAIII with compound heterozygous variants in *RACGAP1*, which encodes CYK4/MgcRacGAP, the other component of the centralspindlin complex.

## Results and Discussion

The proband initially presented as a toddler with unexplained macrocytic anemia that has persisted throughout his life, but was never severe enough to require a transfusion (Supplemental Fig. 1A). Initial testing ruled out common causes of macrocytic anemia including vitamin B12-deficiency, folate-deficiency, and hypothyroidism (data not shown). Red blood cell (RBC) adenosine deaminase activity (ADA), which can be elevated in patients with Diamond-Blackfan anemia (DBA), was also found to be normal (data not shown). A bone marrow biopsy was performed, which showed striking dyserythropoiesis with megaloblastoid changes accompanied by multi-nucleated erythroid precursors and characteristic gigantoblasts (Fig. 1A), thereby establishing the diagnosis of CDAIII. To determine the underlying genetic cause of this patient’s CDAIII, whole exome sequencing (WES) was performed as part of his clinical care. The previously reported p.P916R variant in *KIF23* (7) was not seen in the proband, nor were any other putative causative variants in *KIF23* (data not shown). However, the proband did have compound heterozygous variants in *RACGAP1*, namely, c.1187T>A; p.L396Q (inherited from parent 1) and c.1294C>T; p.P432S (inherited from parent 2) (Fig. 1B). Complete blood counts were performed on the parents, and neither was anemic (Supplemental Fig. 1B), suggesting that these alleles are recessive. Neither variant was seen in the genome aggregation database (gnomAD) (16), indicating that both variants are extremely rare. In addition, both residues are highly conserved through evolution (Fig. 1C) with each alteration resulting in non-conservative amino acid substitutions. Indeed, both variants have a combined annotation dependent depletion (CADD) score of >25 and both are predicted to be deleterious by several other *in silico* algorithms including Provean, PolyPhen, SIFT, and Mutation Taster. These results suggested that the compound heterozygous variants in *RACGAP1* variant identified in this patient may be the first known genetic cause of an autosomal recessive form of CDAIII.

**Figure 1:**
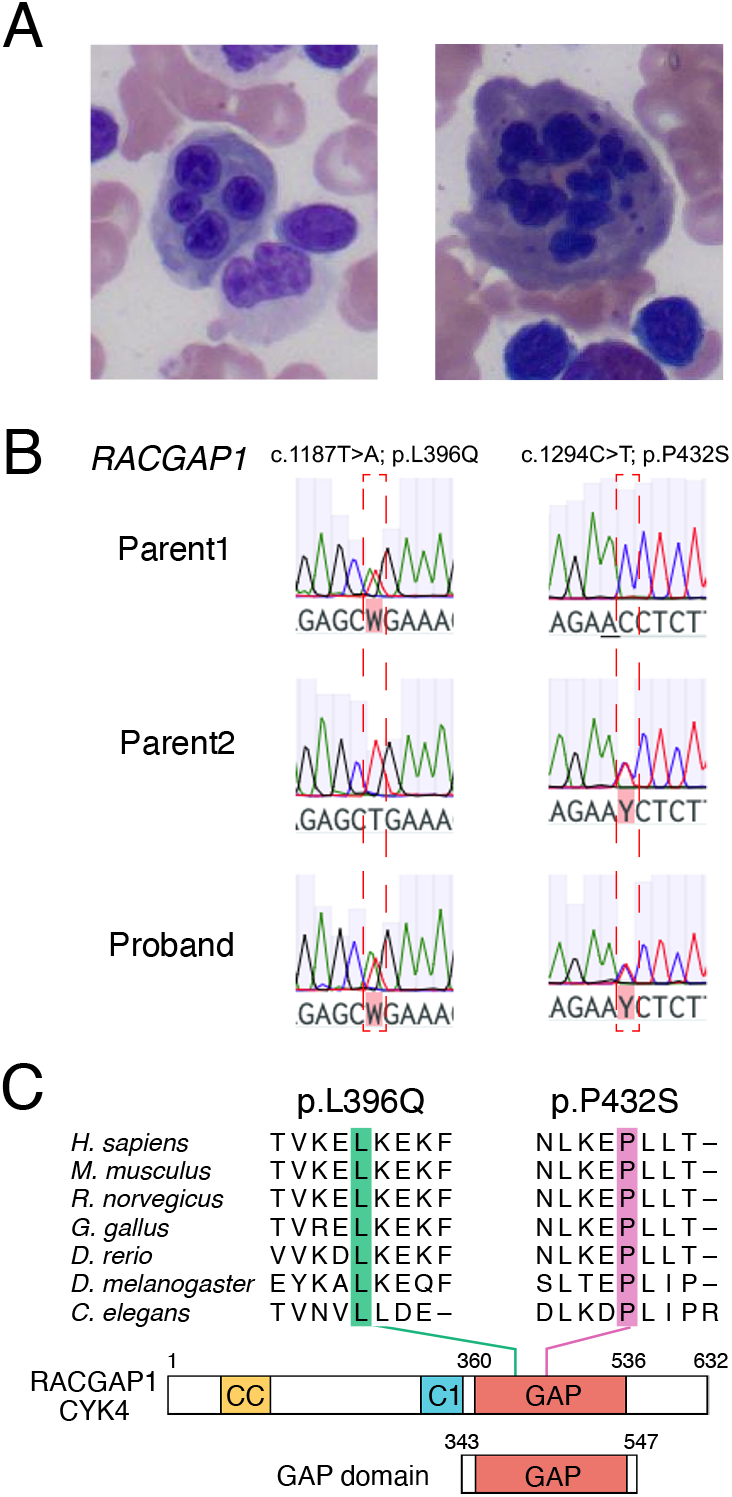
Identification of a patient with CDAIII that has compound heterozygous variants in *RACGAP1*. A) A bone marrow aspirate from the proband showing a characteristic multi-nucleated erythroblast (left) and a gigantoblast (right). B) Electropherograms for the Parent 1 (top), Parent 2 (middle), and the proband (bottom) for variants c.1187T>A (left panels) and c.1294C>T (right panels). C) Domain structure of CYK4 showing the coiled-coil (CC), C1, and RhoGAP (GAP) domains with the MKLP1-interacting region. Sequence alignment of CYK4 orthologs showing the strong conservation of indicated residues. The GAP domain fragment (aa 343 – 547) was used for the biochemical assays in Fig. 3.

To test the functional consequences of these variants, an RNAi-mediated depletion and rescue assay was performed, which is similar to the experiments that established the pathogenicity of p.P916R in *KIF23* (7). At least two independent, stable, HeLa cell lines were generated expressing an RNAi-resistant full-length CYK4-GFP construct (17) containing the wild-type (WT) sequence, the p.L396Q variant, or the p.P432S variant, along with a GFP only control (Fig. 2). Consistent with the findings in this family, we were able to generate stable lines from both variants, indicating that they do not have a strong dominant effect. Immunoblotting revealed that expression levels of the CYK4-GFP transgenes varied between stable lines (Fig. 2A and B) but were comparable to the endogenous protein (Supplementary Figure 2). These cells were then subjected to RNAi to deplete endogenous protein levels and live imaging was performed to determine the effect of each construct on cytokinesis. Cells expressing GFP alone showed a high frequency of cytokinesis failure, resulting in multinucleated cells, whereas this effect was strongly suppressed in cells expressing WT CYK4-GFP (Fig. 2C). Of note, there was a correlation between WT CYK4-GFP levels, which was monitored by GFP fluorescence, and the ability to rescue the cytokinesis defect; this allowed us to establish an expression threshold where WT CYK4-GFP was able to rescue the cytokinesis defect (Fig. 2B). Using this threshold, we find the p.L396Q variant had a modest, but significant, defect in rescuing the phenotype (Fig. 2B and C). One stable line with high expression of this variant (L15 – see Fig. 2A and B) showed a modest inability to rescue cytokinesis, whereas the other stable line (L6) showed a stronger deficit in rescuing the cytokinesis failure (Fig. 2B and C). Strikingly, the p.P432S variant almost completely failed to rescue the depletion of the endogenous CYK4 in all three independent lines, regardless of the transgene expression (Fig. 2B and C). Thus, both variants negatively affect the ability of CYK4 to promote proper cytokinesis.

**Figure 2:**
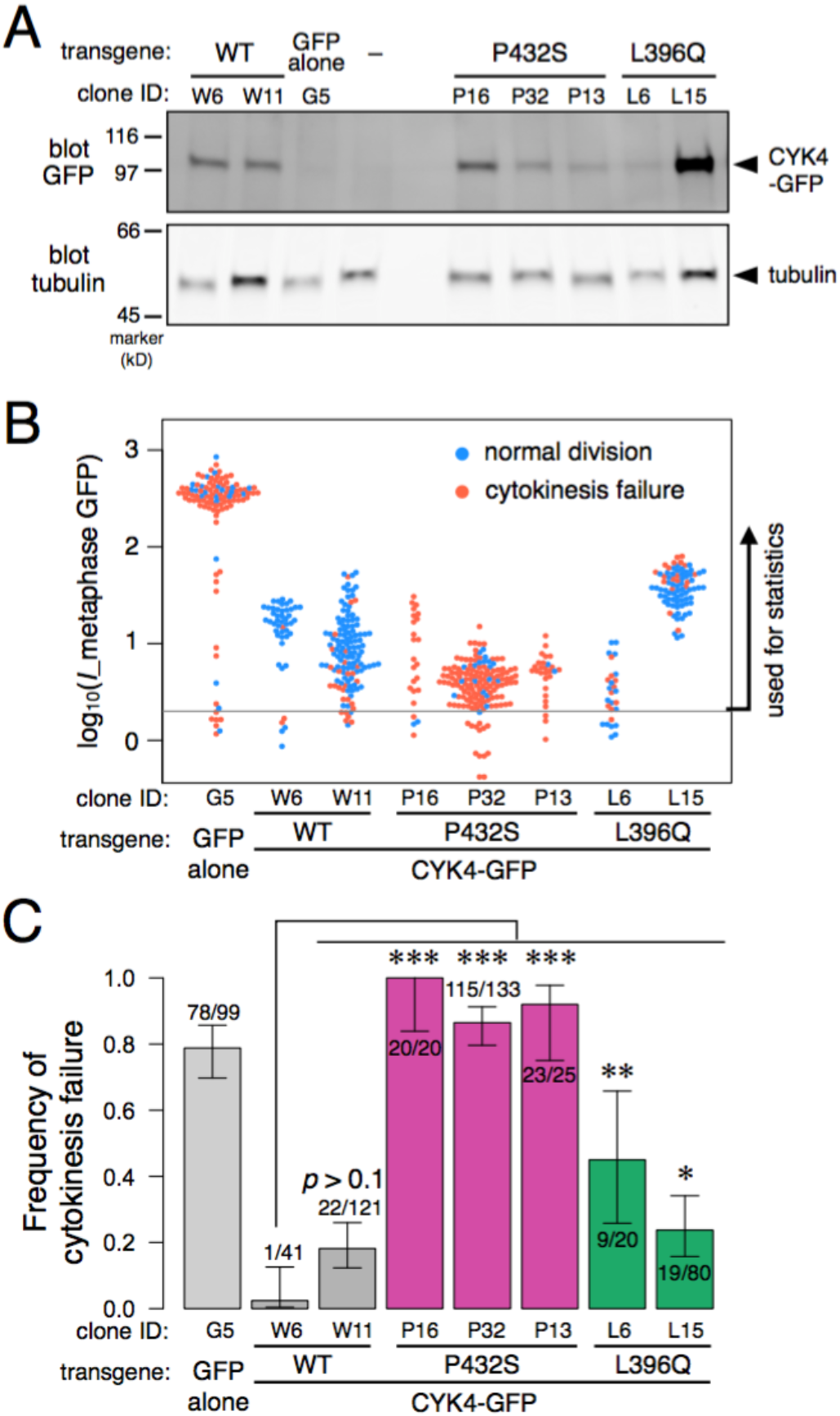
CYK4 variants result in cytokinesis failure. A) Monoclonal HeLa Kyoto cell lines stably transformed with *RACGAP1* cDNA constructs or a control GFP construct were generated and assessed for the expression of CYK4-GFP by immunoblotting mitotic lysates with an anti-GFP antibody (blot GFP). The parental HeLa Kyoto cells (“–”) serve as a negative control. The same membrane was blotted for tubulin as a loading control (blot tubulin). B) The expression level of CYK4-GFP was monitored by the GFP signal during metaphase after siRNA-mediated depletion of the endogenous CYK4. The mean intensity above the background signal of individual cells was plotted on a log scale (dots) with the success or failure of cytokinesis by color. C) The frequencies of cytokinesis failure calculated from the indicated number of cytokinesis failure among the observed cells in at least three independent experiments per line are shown with the 95% confidence interval. The *p*-values (***, *p* < 0.001; **, *p* <0.01; *, p < 0.05) of the statistical test for the difference from the WT cell line (w6) were corrected for multiple comparisons (Dunnett).

We next sought to understand the mechanism by which these variants led to defective cytokinesis. A GAP domain stimulates the GTP hydrolysis by a GTPase, such as RhoA, Cdc42, or Rac1, using a conserved arginine residue (the so called “arginine finger,” R385 in CYK4) (18–20). This facilitates the conversion of the target GTPase from the GTP-bound to the GDP-bound form, which typically results in switching off their downstream effectors (21). The GAP domain of CYK4 stimulates Cdc42 and Rac1 with much greater efficiency than RhoA (22–25). The structural basis for this specificity is unclear since most amino acid residues on the GAP-binding surface of the Rho-family GTPases are conserved among these members (Fig. 3A, light pink). Only three residues closely located to each other near the periphery of the binding surface are substrate-specific (i.e., D90, E97, and M134 in RhoA; S88, E95, N132 in Cdc42, Fig. 3A) (29, 31) and evolutionarily conserved (Supplementary Figure S3). The counterparts in CYK4 are found on a structural motif consisting of two helixes linked by a loop (Fig. 3A, cyan) between R385 (blue) and P432 (magenta), which also includes L396 (green). Thus, we investigated whether these new CDAIII variants might affect the GAP activity of CYK4 towards RhoA, Rac1, and Cdc42.

**Figure 3:**
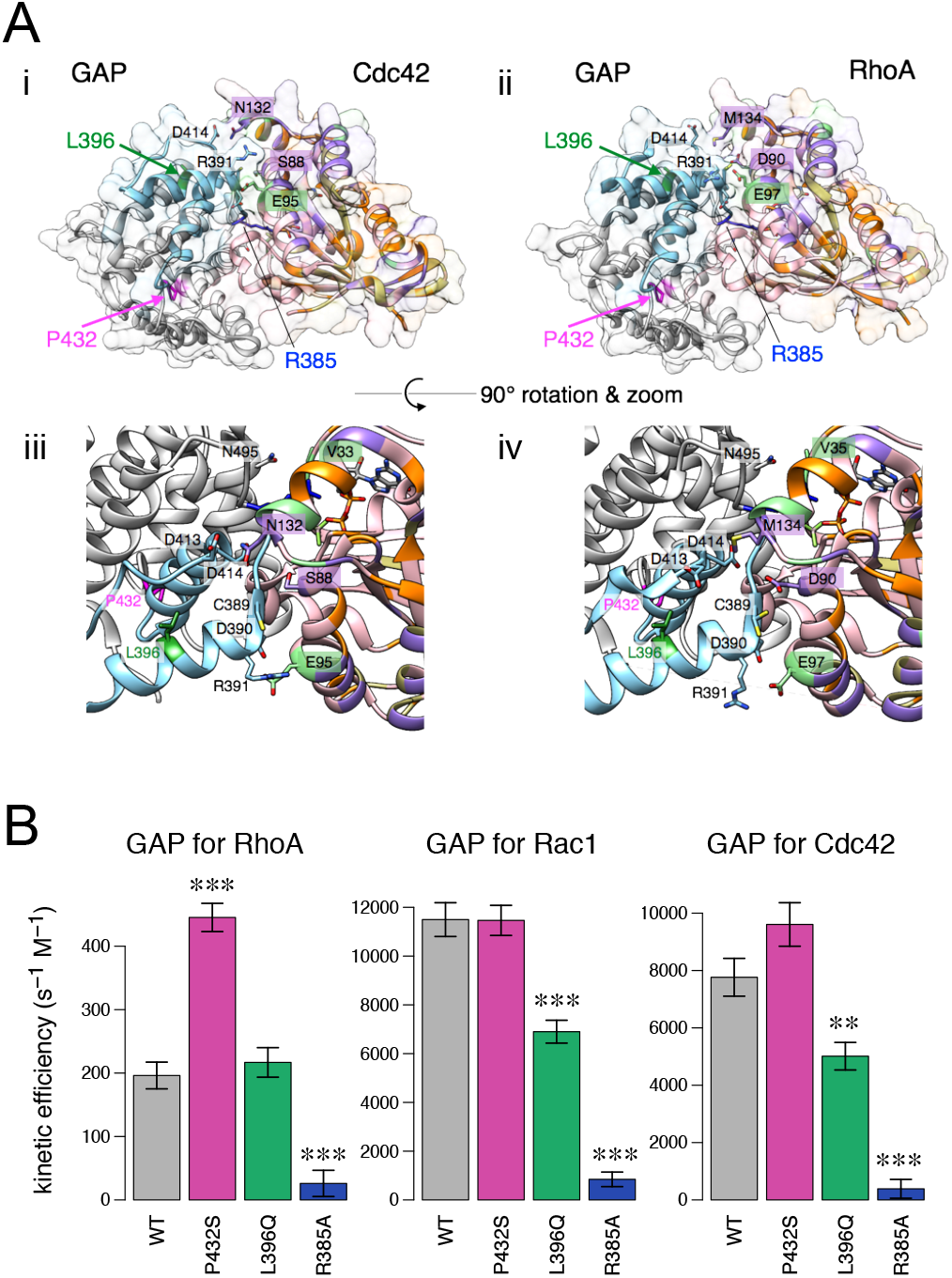
CYK4 variants result in altered GAP activities towards RhoA, Rac1, and Cdc42. A) Crystal structures of the CYK4 GAP domain (left) in a complex with Cdc42 (i and iii, PDB:5C2J) or with RhoA (ii and iv, PDB:5C2K) (right). The residues of the GTPases are color-coded for the difference between the subtypes of Rho GTPases (canonical Rho, Rac, and Cdc42): pink, conserved among the three subtypes; orange, Rho-specific (i.e., identical between Rac and Cdc42); green, Rac-specific; khaki, Cdc42-sepcific; purple, different in all the three. The arginine finger (R385) and the residues found to be mutated in this study (L396 and P432) on the GAP are marked in blue, green and magenta, respectively. The region between R385 (blue) and P432 (cyan) makes contact with the three subtype-specific residues on the GTPases (S88, E95, and N132 on CDC42 and D90, E97, and M134 on RhoA. B) The GAP activity, i.e., the stimulation of GTPase of human RhoA, Cdc42 and Rac, by the CYK4 GAP domain with or without mutations (WT, p.P432S, p.L396Q, p.R385A). The kinetic efficiency (*k*_cat_/*K*_M_) was estimated with a linear model between the rate of GAP-stimulated GTPase activity versus the concentration of the GTPase (Supplementary Figure 4) and shown with the standard error. The *p*-values (***, *p* < 0.001; C) **, *p* <0.01) of the statistical test for the difference from the control (WT GAP) were corrected for multiple comparisons (Dunnett).

GTP hydrolysis by RhoA, Rac1, or Cdc42 was monitored by measuring the released free phosphate using an enzyme coupled photometric assay under a multiple turnover condition in which the GDP release is not rate-limiting (26, 27). The GAP activity was determined as the increase of the GTPase rate in the presence of the CYK4 GAP domain (Fig. 1C). As a negative control, we included a mutant for the ‘arginine finger’ (p.R385A) (24, 28). As expected, WT CYK4 showed significantly stronger ability to stimulate the GTPase activities of Cdc42 and Rac1, as compared to that of RhoA (Fig. 3B). As expected, the p.R385A negative control severely diminished all the GTPase activities, almost to the basal level. Interestingly, the p.P432S variant showed no effect on stimulation of Cdc42 and Rac1, but significantly elevated the ability to stimulate RhoA (Fig. 3A). In contrast, the p.L396Q variant showed no difference in stimulating RhoA activity, but caused a significant decrease in the ability to stimulate Cdc42 and Rac1 activity (Fig. 3B). These results suggest that both variants differentially modify the substrate specificity of the CYK4 GAP domain.

As centralspindlin is a key signaling hub for cytokinesis (29, 30), we also tested whether the variants affected CYK4 localization during cell division. WT-CYK4 showed diffuse cytoplasmic localization during metaphase but began to accumulate at the spindle midzone at the onset of anaphase (Fig. 4A, white arrows), as previously reported (12, 17, 24, 31–33). The p.L396Q variant showed a similar pattern of localization (Fig. 4B). In contrast, the p.P432S variant failed to accumulate sharply at the spindle midzone, but instead localized to the equatorial cell cortex with a ring-like pattern (Fig. 4C, magenta arrows). This abnormal localization was often accompanied by a failure to form a cleavage furrow or by formation of a transient shallow furrow that failed to complete (Fig. 4C). Taken together, these results demonstrate that the severe cytokinesis defect of the p.P432S variant (Fig. 2B and C) is likely due to the elevated GAP activity towards RhoA (Fig. 3B) and severe mislocalization (Fig 4B), whereas the modest cytokinesis defect of the p.L396Q variant (Fig. 2B&C) is primarily due to the diminished GAP activity towards Rac and Cdc42 (Fig. 2B).

**Figure 4:**
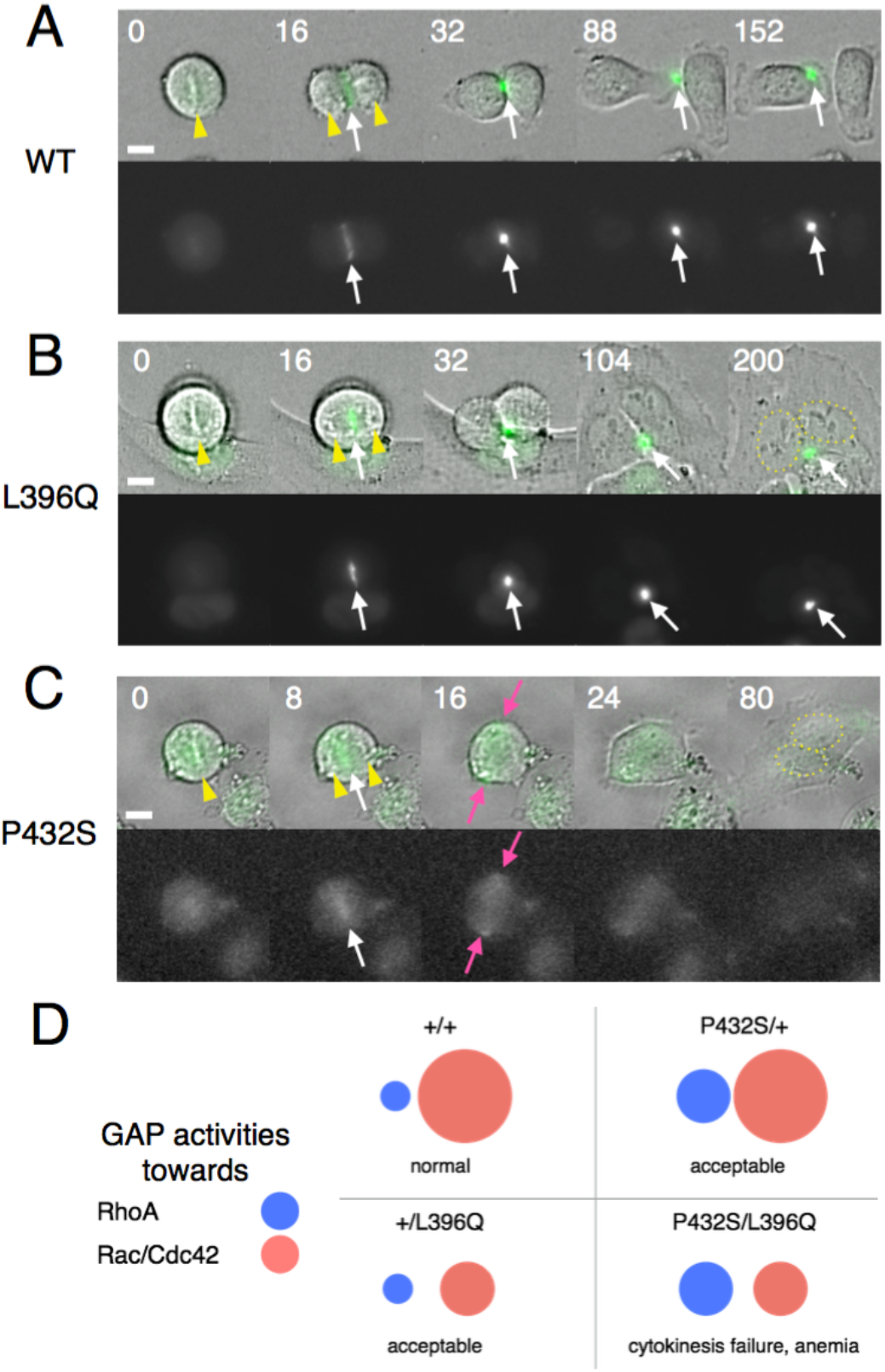
CYK4 variants cause distinct cytokinesis phenotypes. A-C) Stills from live microscopy of cells stably expressing the wildtype (WT) or a variant CYK4 tagged with GFP after depletion of the endogenous protein (top, bright field and GFP in green; bottom, GFP in grayscale). The transition from metaphase to anaphase was monitored by chromosomes (yellow arrowheads). White arrows indicate the localization of each of the indicated constructs. D) Proposed model for the significance of target preference of the GAP domain of CYK4 in cytokinesis. The elevated activity of p.P432S on RhoA in combination with the reduced activity on Rac/Cdc42 by the p.L396Q variant disturbs the proper balance between these Rho GTPases important for cytokinesis.

Most reported CDAIII cases derive from studies of two families with a dominant missense mutation in *KIF23* (7). However, reports of sporadic cases suggested the existence of autosomal recessive form(s) with unknown genetic etiology (1, 2, 34, 35). Here we report a novel sporadic case of CDAIII and identify compound heterozygous variants in *RACGAP1* (Fig. 1), which forms the centralspindlin complex with *KIF23*. Severe loss-of-function (LoF) variants in *RACGAP1* are very common within gnomAD (as indicated by a probability of LoF intolerant (pLI) score of 0), whereas such LoF variants are rarely seen in *KIF23* (pLI score of 1) (16). This suggests that strong LoF variants in *RACGAP1* are likely to be recessive, whereas such variants would be dominant in *KIF23* due to haploinsufficiency. Neither of the two *RACGAP1* variants in the proband showed strong dominant negative effects, but both result in defects in cytokinesis in the absence of the wild-type protein (Fig. 2). This is at least partially due to altered GAP activity of both variants (Fig. 3) along with mislocalization of the p.P432S variant (Fig. 4).

The role of the GAP domain in CYK4 has long been an enigma (36–38). Our findings demonstrate the importance of proper substrate specificity of the CYK4 GAP domain for successful cytokinesis. The cortical mislocalization of the p.P432S variant (Fig. 4), which exhibits an elevated GAP activity against RhoA but not to Rac1 or Cdc42 (Fig.3), indicates the importance of the proper target specificity of the GAP domain for the subcellular localization of centralspindlin. This implies a novel mechanism for the cortical recruitment of centralspindlin via the RhoA-GAP interaction, which is hidden during the normal cell division, but might contribute to the cleavage signalling when the central spindle is disrupted (39, 40). We propose a model based on the GAP activity that tries to explain the recessive nature of the identified variants (Fig. 4B).

An important question that still remains is why dysfunction of the centralspindlin complex, which is broadly expressed, leads to a predominately RBC-restricted phenotype. This is reminiscent of the ribosomal protein gene mutations associated with DBA, which is another form of congenital macrocytic anemia. Similar to DBA (41), CDAIII may serve as an additional model to understand how cell-type restricted phenotypes arise from broadly expressed genes.

In conclusion, our findings strongly suggest that CDAIII is a disease of abnormal centralspindlin complex function. Mutations in other proteins that interact and function with centralspindlin (42) may also lead to a similar phenotypes in humans. Indeed, viewing diseases as aberrancies of genetic pathways versus defects in individual genes may provide the proper framework on which to build a taxonomical structure of disease.

## Methods

See supplemental methods for more detailed information.

### Study approval

The Columbia University Irving Medical Center (CUIMC) IRB panel deemed this study exempt from IRB approval because it was considered a case study.

## Data Availability

Requests for any data presented can be obtained through contacting the corresponding authors.

## Author contributions

SNW – acquiring data, analyzing data, providing reagents, and writing the manuscript

MB – conducting experiments, acquiring data, analyzing data, providing reagents

ST, BHD, ML – acquiring data

MM – designing research studies, analyzing data, and writing the manuscript

## Acknowledgments

We thank the patient and his parents for their cooperation throughout the studies. We thank Drs. Sergey Lekomtsev and Mark Petronczki for plasmids, and Drs. Vimla Aggarwal, Julie Canman, Kartik Ganapathi, Kazutaka Murayama, Mikako Shirouzu, Steve Spitalnik, and Jenny Yang for helpful discussions and/or critical comments on the manuscript. This work was supported by 1K08NS119567 to SNW and a Cancer Research UK program grant (C19769/ A11985) to MM.

