## Supplemental for "A rare congenital anemia is caused by mutations in the centralspindlin complex"

**Supplemental Methods**

*Whole exome-sequencing:* Clinical whole exome sequencing was performed on the proband and mother in the CLIA- and CAP-certified laboratory of Personalized Genomic Medicine (PGM) at CUIMC as part of the patient’s routine care. Di-deoxy sequencing was performed in PGM from the proband and parents to determine the phasing of the variants.

*Generation of cell lines*: The monoclonal HeLa Kyoto cell lines stably expressing the variants of CYK4-GFP were generated as previously described (1). The c.1294C>T and c.1187T>A variants were introduced by site-directed mutagenesis into the pIRESpuro3-RACGAP1-FLAc plasmid, which encodes the full-length CYK4 resistant to an siRNA targeting the endogenous *RACGAP1* (Invitrogen Stealth HSS120934, GCCAAGAACUGAGACAGACAGUGUG). Transgene expression in bulk populations was assessed by immunoblotting of the crude lysate prepared from nocodazole-arrested mitotic cells with a mouse monoclonal anti-AcGFP antibody (JL8, Clontech) or a mouse monoclonal anti-CYK4 antibody (1G6, Abnova).

*Live microscopy*: For live microscopy, the cell lines were cultured in a 35-mm glass-bottom dish (Fluorodish, FD35, World Precision), transfected with the above siRNA using Lipofectamine RNAiMAX (Invitrogen), incubated for 12 h, and treated with 2.5 mM thymidine for 16 h. Recording of the differential interference contrast (DIC) and GFP images was started 8 h after release from the thymidine block using a DeltaVision system (Applied Precision) as described previously (2). The expression level of CYK4-GFP in individual cells was assessed by the mean intensity of the whole cytoplasm during metaphase.

*GAP activity assay*: The cDNA fragments of the CYK4 GAP domain (aa residues 343-547) with or without the mutations were amplified with a primer pair (ACTGCCATGGGACCTGTCAAGATTGGAGAGGGAATG and AGTCCTCGAGTCATTGCTCCACCATCATGAACTGAC) and subcloned into the NcoI/XhoI site of pGEX-TEV vector. The recombinant proteins expressed in the BL21(DE3) *E. coli* cells were bound to glutathione-Sepharose 4B (GE Healthcare), eluted by cleavage with AcTEV protease (Invitrogen), and further purified by ion exchange and size exclusion chromatography (Mono Q and Superdex 200, respectively) on an Äkta FPLC system (GE Healthcare). The GAP activity of these preparations was determined as the increase of the GTPase activity of RhoA, Rac1, or CDC42 (Cytoskeleton) from the basal level. Phosphate release in a reaction buffer (50 mM HEPES pH7.6, 100 mM NaCl, 0.95 mM EDTA, 0.2 mM GTP) was monitored with EnzChek Phosphate Assay Kit (Invitrogen) in a 384-well plate (Corning) by a Varioskan Flash plate reader (Thermo Scientific) at 22 °C.

**A**

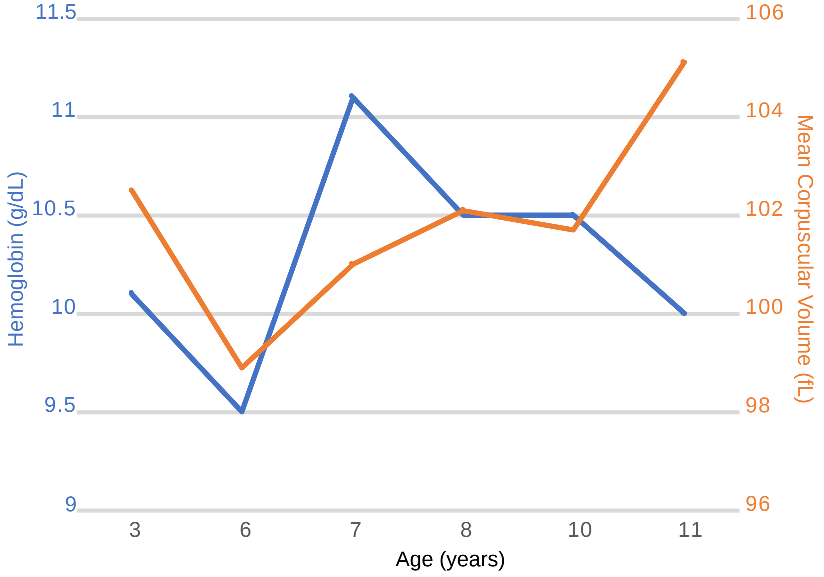


**B**

| | Parent 1 | Parent 1 | Parent 2 | Parent 2 |
|---|---|---|---|---|
| HGB (g/dL) | 13.4 | 13.4 | 15.1 | 14.5 |
| HCT (%) | 40.3% | 39.8% | 45.5% | 43.8% |
| MCV (fL) | 89.6 | 86.9 | 98.7 | 94.4 |

**Supplemental Figure 1: The proband has a chronic macrocytic anemia but parents are not anemic**

A) Hemoglobin and the mean corpuscular volume (MCV) are plotted as a function of the proband's age in years. The blue line represents the detected level of hemoglobin (g/dL) as indicated by the left ordinate. The red line represents the measured MCV (fL), the value of which is shown in the right ordinate.

B) Two independent CBCs were performed on the proband's parents with the relevant RBC indices shown.

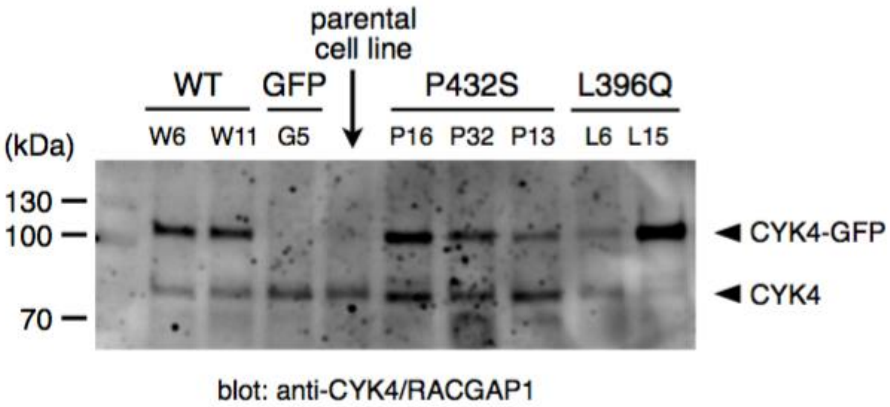


**Supplementary Figure S2: The expression levels of the CYK4-GFP transgene is comparable to that of the endogenous CYK4.**

Mitotic cell lysates from the indicated cell lines were analyzed by immunoblotting with anti-CYK4 antibody (1G6, Abnova), which detects both the endogenous CYK4 and the CYK4-GFP transgene.

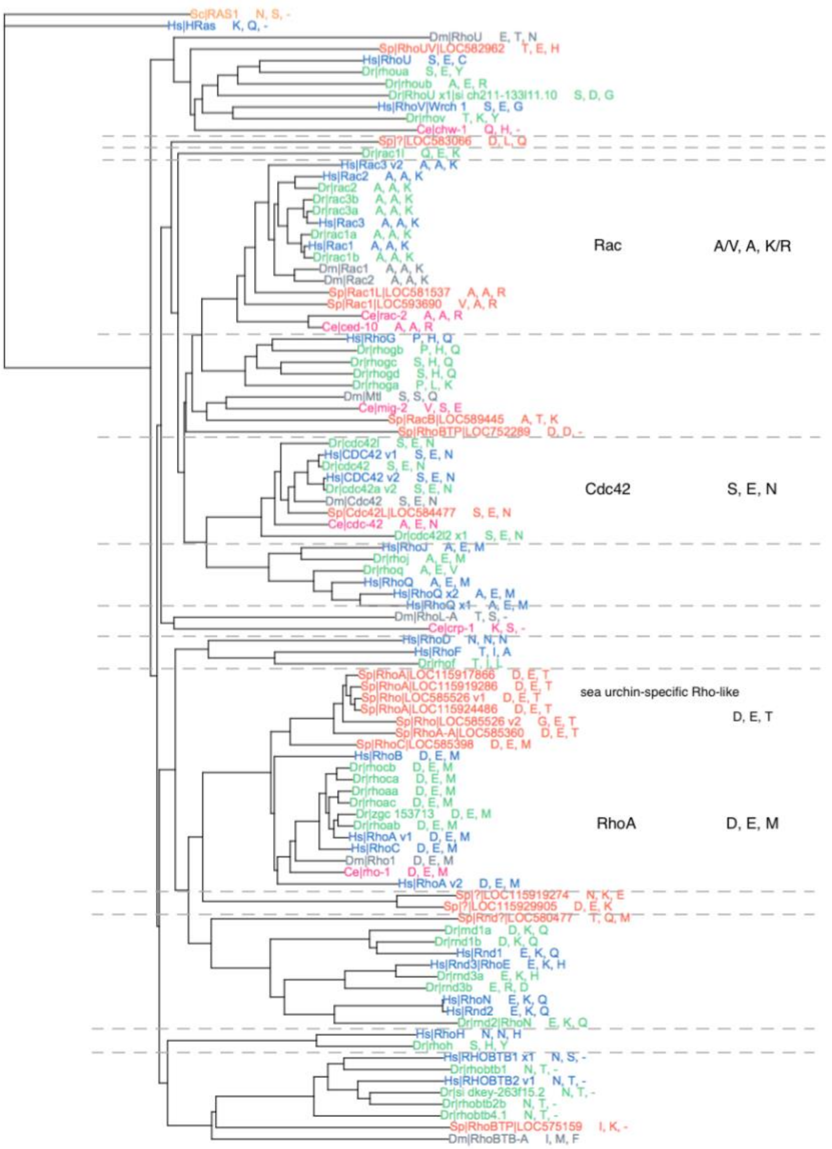


**Supplementary Figure S3: The subtype-specific residues on the GAP-interacting surface of the Rho-family small GTPases are evolutionarily conserved.**

A phylogenetic tree of the Rho-family small GTPases from human (*Homo sapiens*, Hs, blue), zebrafish (*Danio rerio*, Dr, green), see urchin (*Strongylocentrotus purpuratus*, Sp, orange) fruit fly (*Drosophila melanogaster*, Dm, gray), and nematode (*Caenorhabditis elegans*, Ce, magenta) with human and yeast Ras proteins as outgroups. The three variable residues on the GAP-interacting surface, which correspond to D90, E97, and M134 on human RhoA, are shown.

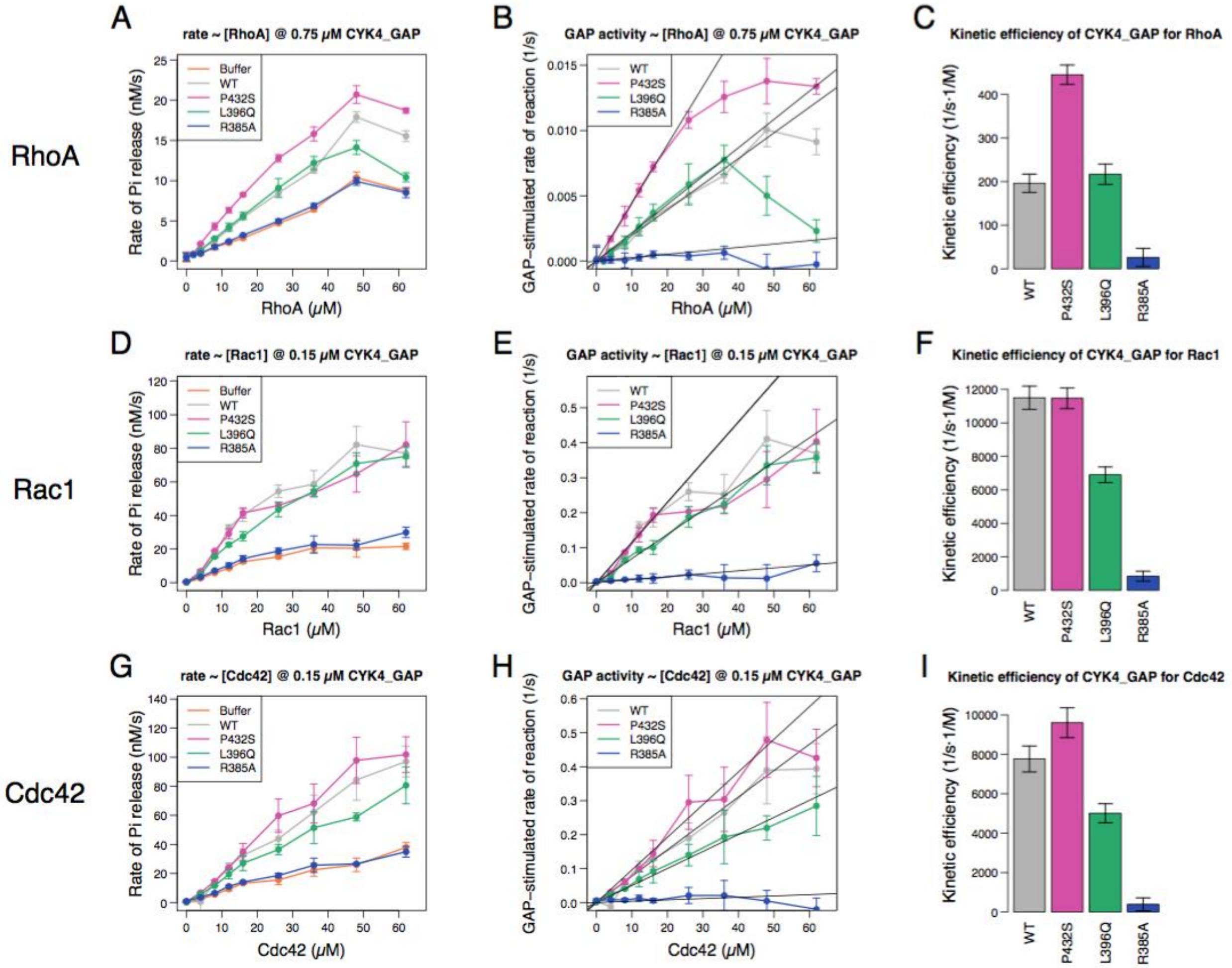


**Supplementary Figure S4: Determination of the kinetic efficiency of the GAP activities of the CYK4 GAP domain towards human RhoA, Rac1 and Cdc42**

A), D) and G) Rate of free phosphate by hydrolysis of GTP by the indicated GTPase in the presence or absence of the CYK4 GAP domain with or without a mutation.

B), E), and H) The rate of catalysis of the reaction by the GAP was determined as the stimulated rate of the GTPase activity per molecule of the GAP.

C), F), and I) The kinetic efficiency ($k_{cat}/K_M$) was estimated as the gradient of the catalytic rate vs GAP concentration curve (B, E, and H) near the origin by linear fitting, and shown with the standard error.
